# Intracranial-EEG Based Classification and Localization of Epileptiform Activity in a Large Cohort of Adults with Drug-Resistant Epilepsy

**DOI:** 10.64898/2025.11.30.25340505

**Authors:** Samantha Yost, Justin M. Campbell, Matthew Findlay, Wendy Sun, Kimble Mahler, Silvia Soule, Coby Soule, Shervin Rahimpour, Ben Shofty

**Affiliations:** Spencer Fox Eccles School of Medicine, University of Utah, Salt Lake City, UT, USA; Department of Neurosurgery, University of Utah, Salt Lake City, UT, USA; Department of Neurology, University of Utah, Salt Lake City, UT, USA

## Abstract

**Objective:** To characterize the type and distribution of epileptiform activity across anatomical regions and functional networks in a large cohort of patients with drug-resistant epilepsy (DRE) undergoing intracranial electroencephalography (iEEG).

**Methods:** We retrospectively reviewed iEEG recordings from 93 patients with DRE who underwent stereoelectroencephalography at our institution between 2019 and 2025. Epileptiform activity was classified as “ictal”, “interictal”, or “ictal spread” based on visual inspection and clinical correlation. Electrode coordinates were localized to anatomical regions using the *Brainnetome Atlas* and to functional networks using the *DU15NET-Consensus Atlas*. The distribution of epileptiform activity across regions and networks was compared with global baselines using Chi-square tests with false discovery rate correction.

**Results:** Interictal discharges were the most prevalent type of epileptiform activity (median 9.5% of contacts per patient), followed by ictal (6.0%) and ictal spread (2.1%). Temporal regions exhibited an increased prevalence of all types of epileptiform activity compared to the global baseline, whereas frontal regions showed marked reductions, despite dense sampling. At the network level, epileptiform activity was overrepresented in Default Network-A and underrepresented in the Salience/Parietal Memory Network.

**Significance:** Epileptiform activity shows consistent, non-uniform patterns across both anatomical regions and functional networks. These findings reinforce the central role of the temporal lobe and associative networks in epileptogenesis and support the view of epilepsy as a disorder of distributed brain networks. Mapping epileptiform activity in a network framework may enhance biomarker development and inform circuit-based surgical and neuromodulatory treatment strategies.

## Introduction

Epilepsy is one of the most common and disabling neurological disorders worldwide.^1^ Moreover, one-third of individuals with epilepsy do not receive adequate relief despite multiple trials of antiseizure medications and are termed drug-resistant (DRE).^2– 4^ A diagnosis of focal, non-lesional DRE is associated with considerable functional impairment—increased medical comorbidities, a heightened risk for injury and death, and significant reductions in quality of life.^5,6^ For these individuals, neurosurgical interventions aimed at either resecting pathological seizure foci or disrupting seizure spread via neuromodulation (e.g., responsive neurostimulation), can be highly efficacious.^7,8^

Intracranial electroencephalography (iEEG) recordings are an invaluable tool for detecting seizure foci and guiding clinical decision-making in epilepsy surgery.^9,10^ These methods facilitate localization of seizure foci via both the passive recording of abnormal neurophysiological signals and active perturbation with direct electrical stimulation.^11,12^ Together these methods have characterized a broad range of epileptiform activity, including interictal epileptiform discharges (IEDs), high-frequency oscillations, and distinct ictal onset patterns.^13^ These varied patterns of activity have been implicated in different aspects of epilepsy pathophysiology and provide insight into the potential pathways of seizure propagation.^14–16^

Though mesial temporal lobe epilepsy is the most common type of adult DRE^17^, recent work has increasingly emphasized that epilepsy is not solely a focal pathology, but instead, involves distributed brain networks.^18–21^ Numerous abnormalities in structural and functional brain networks have been observed in those with epilepsy.^22^ Ictal symptoms, like cognitive impairment and memory dysfunction, for instance, have been linked to these abnormalities in network function caused by epileptiform activity (e.g., IEDs).^23–25^ Nevertheless, most prior investigations of epileptiform activity have been limited by small sample sizes, heterogeneous methodologies, or a narrow focus on single event types or anatomical regions. As a result, there is an incomplete understanding of the prevalence of these types of epileptiform activity across different brain regions and functional networks. Even with the most effective surgical interventions, such as anterior temporal lobectomy (ATL), and initial seizure freedom, many patients experience recurrence over time, likely due to network-level progression and a kindling phenomenon. This underscores the importance of understanding the underlying epileptic networks to improve long-term outcomes and guide future therapeutic strategies.

To address this knowledge gap, we characterized both the type and distribution of epileptiform activity observed across a large cohort of individuals with DRE. We hypothesized that, in line with prior observations, certain regions would emerge as hubs of pathological activity (e.g., mesial temporal lobe). Moreover, we hypothesized that investigating the presence of epileptiform activity across functional networks may provide complementary information about distributed epileptogenic processes beyond what is detectable with focal, region-centric approaches.

## Methods

### Data Collection and Curation

We performed a retrospective review of all patients with DRE who underwent phase 2 intracranial monitoring with stereoelectroencephalography (SEEG) at our institution from 01/2019 through 01/2025 as part of a clinical evaluation to identify seizure foci. This study was approved by the University of Utah Institutional Review Board with a waiver of patient consent. The number, type, and distribution of SEEG electrode contacts varied due to heterogeneity in seizure semiology and anatomical considerations (e.g., presence of focal dysplasia or sclerosis) across individuals.

Demographic information (e.g., age, sex) and clinical characteristics (e.g., number of electrode contacts, subsequent surgery) were obtained by reviewing each patient’s medical record. The presence or absence of epileptiform activity on each contact was noted by a clinical epileptologist examining the continuous iEEG obtained during each patient’s inpatient hospitalization; epileptiform activity was designated as either “ictal” (involved in seizure), “interictal” (abnormal activity present between seizures), or “ictal spread” (contributing to seizure propagation).

### Electrode Localization

To localize electrodes, we used open-source software developed by our group (*Locate electrodes Graphical User Interface, LeGUI*).^26^ Briefly, the preoperative MRI and postoperative CT were co-registered, several image processing steps were implemented (e.g., white matter and grey matter segmentation, correction for postoperative brain shift), 2D and 3D brain models were generated, and electrodes were automatically detected using the artifact present in the postoperative CT. The centroid position for each electrode was calculated in both patient space and MNI space to facilitate group-level comparisons. Anatomical labels were then assigned using the *Brainnetome* atlas^27^, and grouped into six broader regions to simplify analysis: frontal, temporal, parietal, occipital, insular, and subcortical (e.g., thalamus).

### Volumetric Network Analysis & Cortical Surface Visualization

For each subject, electrode contact coordinates in MNI152 1mm space were assigned a functional network label, based on the *DU15NET-Consensus Atlas*.^28^ This atlas is a novel 15-network parcellation derived from intensively sampled fMRI data on 15 individuals (each scanned 8-11 times). Each contact’s MNI coordinates were localized to the closest voxel in the MNI152 1 mm brain volume. Then, a cube-shaped neighborhood of all voxels within a 3 mm radius was sampled, and the most common (modal) network label within this neighborhood was identified. In cases where the modal label remained unassigned/unknown (indicating proximity to white matter or near the edges of the brain), we identified the second most frequent label within the neighborhood; if this secondary label accounted for ≥ 10% of all voxels in the neighborhood, we reassigned the contact to this label; this approach ensured that contacts were not systematically classified as unknown when a clear alternative label was present.

Next, electrode contact coordinates in MNI152 space were used to create spatial region-of-interest masks corresponding to ictal, interictal, and ictal spread contacts. Each contact’s MNI coordinates were localized to the closest voxel in the MNI152 1 mm brain volume. Each contact voxel was then dilated by 3 voxels in all directions using morphological dilation (i.e., forming a cube-shaped neighborhood). Contacts within the same epileptic event type were combined to produce binary masks for each event type and subject. These volumetric masks were projected to the *fsaverage6* cortical surface using the *CBIG Registration Fusion* pipeline.^29^ Projected surface data were binarized for each subject and summed across subjects for each type of epileptiform activity to create group-level maps on the *fsaverage6* cortical surface.

### Statistical Analysis

Demographic and clinical variables are summarized using the appropriate descriptive statistics. To assess whether the distribution of activity types differed across regions and networks, we first performed a global Chi-square test of independence on a contingency table of region (6 levels) or network (15 levels) by epileptiform activity type (ictal, interictal, ictal spread); this test evaluated whether the overall distribution of activity types varied across networks. We then conducted follow-up Chi-square goodness-of-fit tests separately for each region and network. For these tests, the observed counts of activity types were compared to the expected counts calculated from the global proportions across all electrodes. To account for multiple comparisons, *p*-values were adjusted using the Benjamini–Hochberg false discovery rate (FDR) procedure. Results are reported as the difference between the observed proportions and expected proportions under the global baseline (expressed as a percentage of the expected proportion), test statistics, and the FDR-adjusted *p*-values. All analyses were performed using custom Python scripts and open-source software libraries (i.e., *SciPy*^30^, *statsmodels*^31^).

## Results

### Demographics and Clinical Characteristics

We identified 93 eligible patients who underwent phase 2 intracranial monitoring from 01/2019-01/2025. The mean (± SD) age of the cohort was 36.5 (± 11.5) years. Approximately half of the cohort were females (51.6%), and most individuals were White (91.4%). Most patients had intracranial recordings that spanned both hemispheres (77.4%). Across individuals, the mean (± SD) number of recorded electrode contacts was 125.9 (± 39.2); range 50-229. Following phase 2 monitoring, most patients underwent definitive epilepsy surgery; neuromodulatory interventions (e.g., responsive neurostimulation) were most common (47.3%), followed by resection (20.4%), and ablative procedures (14.0%). A minority of patients (5.4%) received multiple therapies (e.g., ablation followed by neuromodulation), while 12.9% had not yet undergone definitive surgery at the time of data analysis for this report. A summary of the demographics and clinical characteristics is reported in **Table 1**.

**Table 1.** Demographic information and clinical characteristics of the study cohort.

| Variable |  |
| --- | --- |
| Age (yrs., mean ± SD) | 36.5 (± 11.5) |
| fSex (no. female, %) | 48 (51.6%) |
| Ethnicity (no., %) |  |
| White | 85 (91.4%) |
| Hispanic or Latino | 4 (4.3%) |
| Black | 1 (1.1%) |
| American Indian or Alaska Native | 1 (1.1%) |
| Unknown or Unspecified | 1 (1.1%) |
| Bilateral coverage (no., %) | 72 (77.4%) |
| Electrode contacts (mean ± SD) | 125.9 (± 39.2) |
| Subsequent surgery (no., %) |  |
| Neuromodulation | 44 (47.3%) |
| Resection | 19 (20.4%) |
| Ablation | 13 (14.0%) |
| Combined | 5 (5.4%) |
| None Performed | 12 (12.9%) |

### Types and Distribution of Epileptiform Activity

Epileptiform activity recorded from intracranial electrodes was classified as either ictal, interictal, or ictal spread. Ictal spread being defined as a measurement of propagation of seizure activity downstream from the initial active site in the brain. Most patients exhibited each type of activity: ictal (90.3%), interictal (95.7%), and ictal spread (58.1%) (**Figure 1A**). However, the percentage of electrode contacts with epileptiform activity observed in each patient differed across types. Interictal activity was most widely observed (median 9.5% electrode contacts), followed by ictal activity (6.0%) and ictal spread activity (2.1%). Kolmogorov-Smirnov tests comparing the distributions of electrode contacts with each type of activity revealed that interictal activity was more prevalent than ictal activity (*D* = 0.25, *p* = 0.007) and ictal spread activity (*D* = 0.43, *p* < 0.001), and ictal activity was more prevalent than ictal spread activity (*D* = 0.39, *p* < 0.001) (**Figure 1B**).

**Figure 1.**
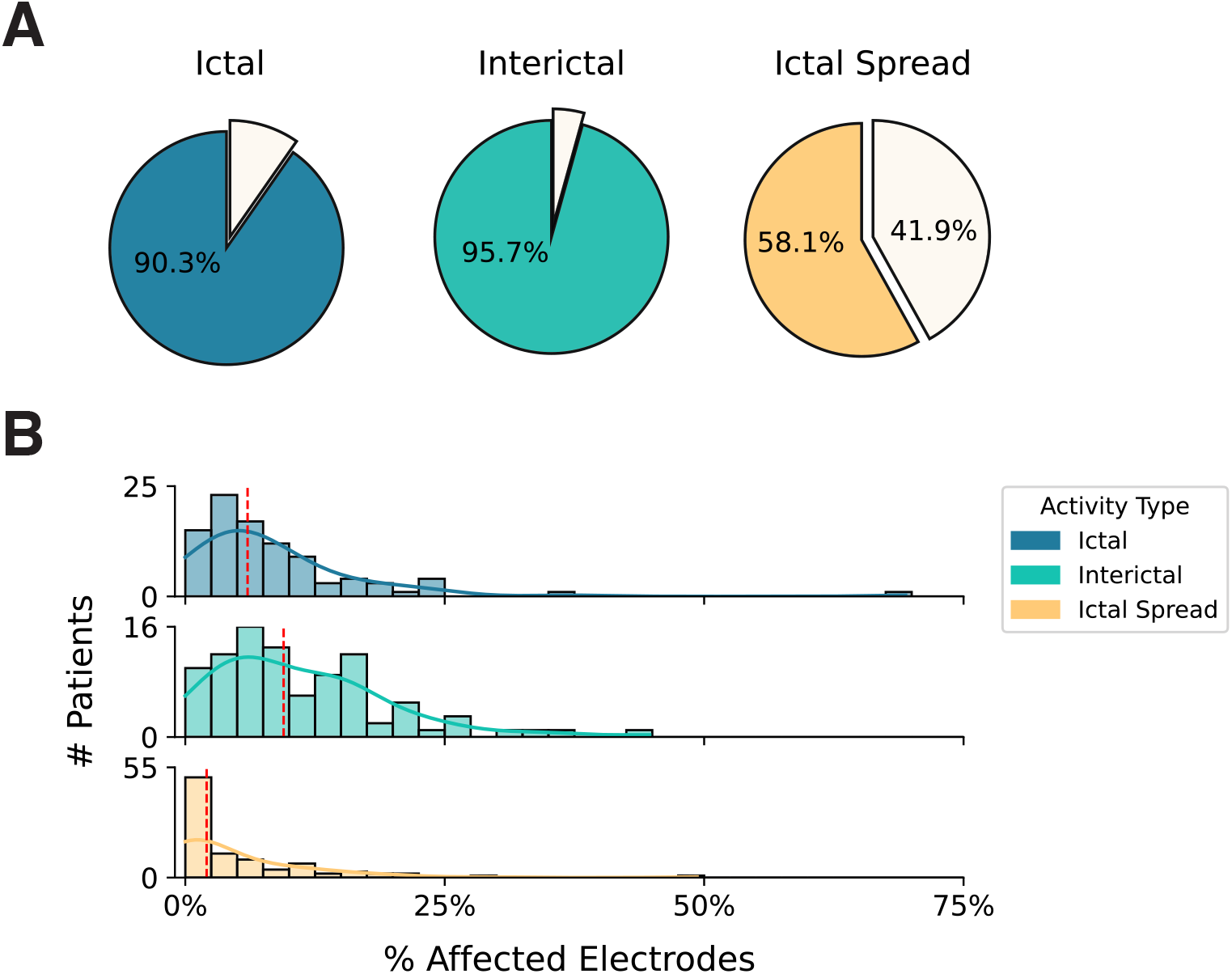
Summary of detected epileptiform activity and the proportions of affected electrodes. (A) The percent of patients in the study cohort with detected ictal (blue), interictal (green), and ictal spread (orange) activity. (B) Histograms of the percent of affected electrodes for each patient, separated by type of epileptiform activity. Solid lines depict the smoothed, Gaussian kernel density probability estimate. Red dashed lines indicate the median percent of affected electrodes across patients. Kolmogorov-Smirnov tests were performed to characterize significant differences in the distributions of epileptiform activity.

Using conventional anatomical labels, we observed that the most densely sampled targets for SEEG were temporal (n = 3,683) and frontal regions (n = 2,047). The most sampled functional network was DN-B (Default Network-B; n = 1,622), which involves distributed brain regions (e.g., lateral temporal cortex, dorsomedial prefrontal cortex) (**Figure 2A)**.

**Figure 2.**
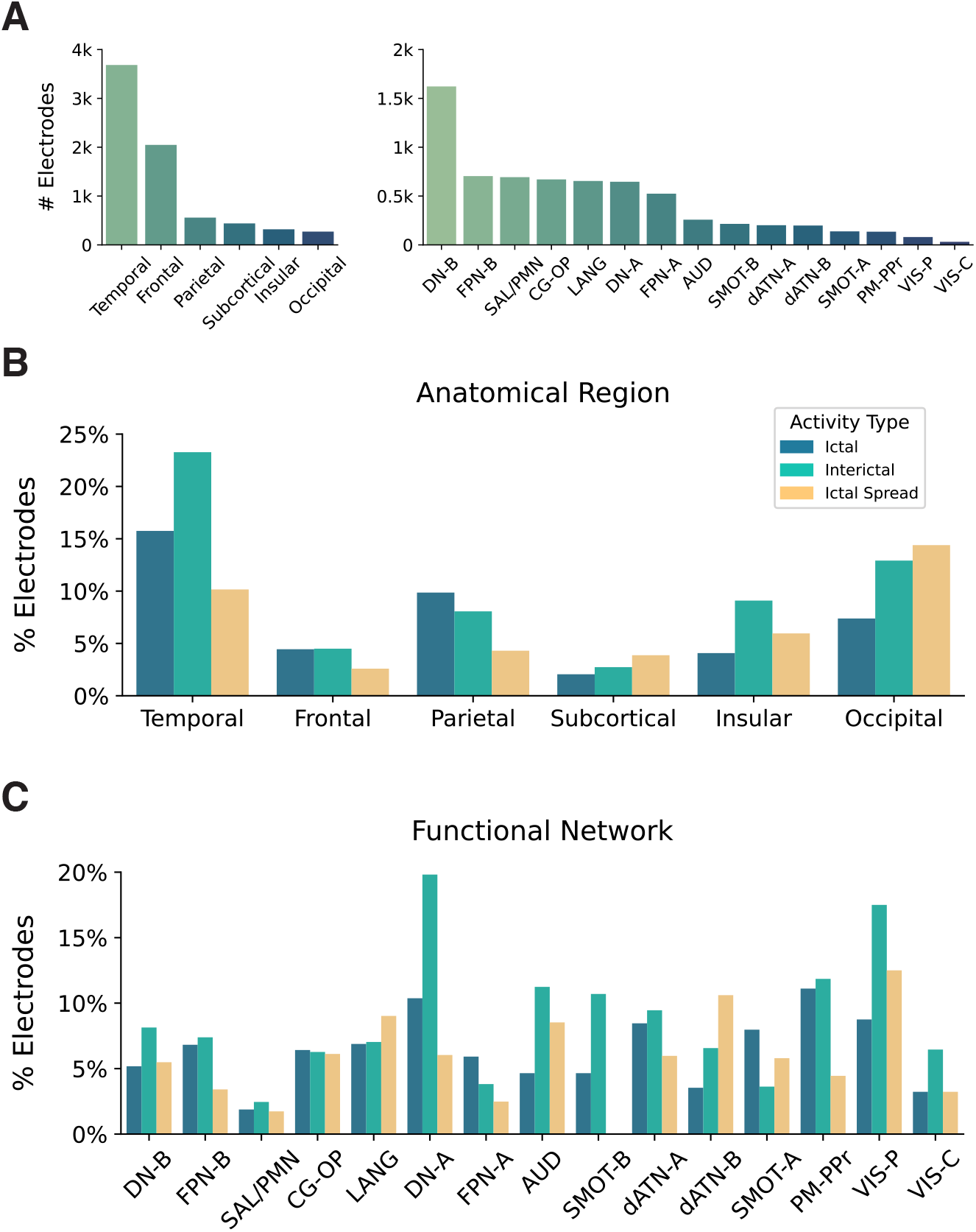
Distribution of epileptiform activity across anatomical regions and functional networks. (A) The total number of electrodes within each anatomical region and functional network. (B) The percent of electrodes with each type of epileptiform activity, grouped by anatomical region. (C) The percent of electrodes with each type of epileptiform activity, grouped by functional network. DN-B = Default Network-B, FPN-B = Frontoparietal Network-B, SAL/PMN = Salience / Parietal Memory Network, CG-OP = Cingulo-Opercular, LANG = Language, DN-A = Default Network-A, FPN-A = Frontoparietal Network-A, AUD = Auditory, SMOT-B = Somatomotor-B, dATN-A = Dorsal Attention-A, dATN-B = Dorsal Attention-B, SMOT-A = Somatomotor-A, PM-PPr = Premotor-Posterior Parietal Rostral, VIS-P = Visual Peripheral, VIS-C = Visual Central.

**Figure 3.**
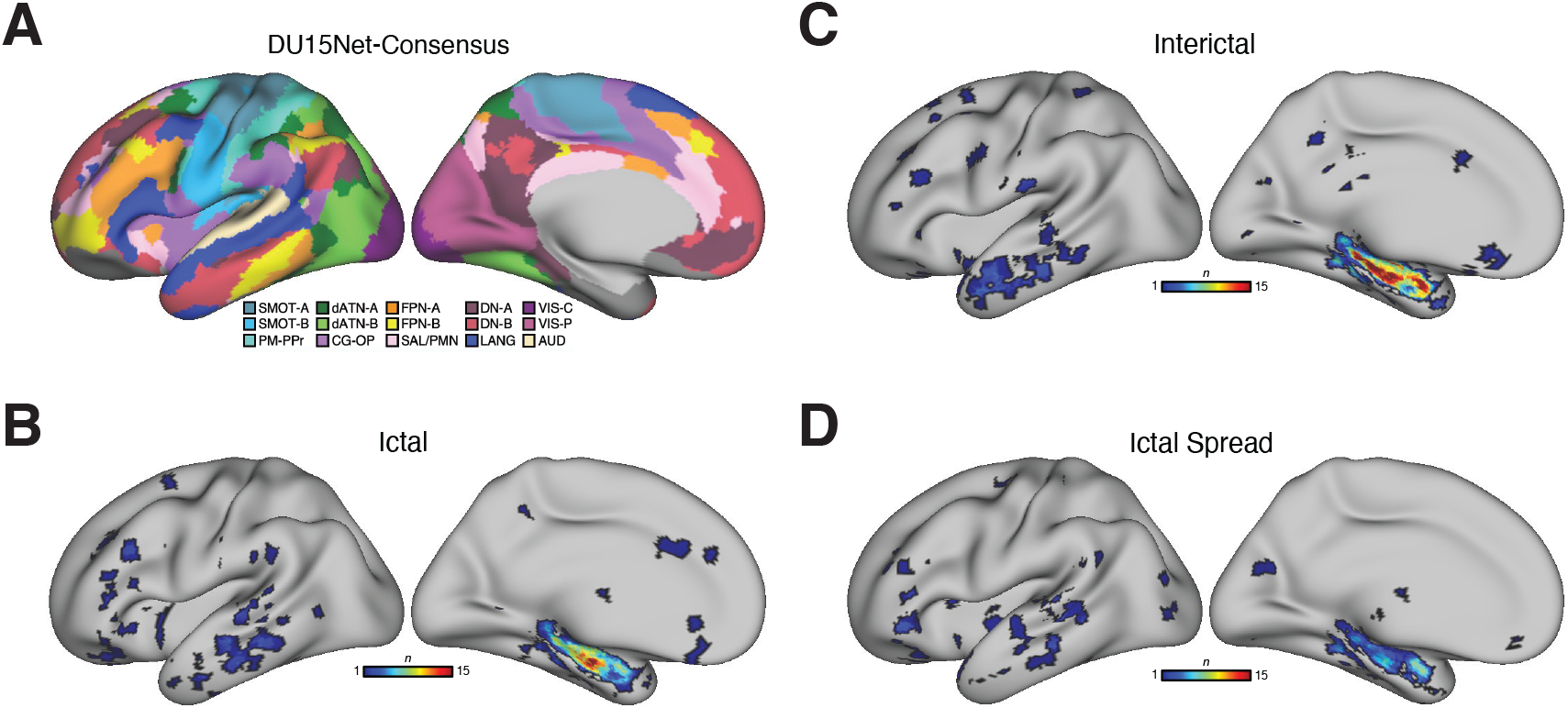
Cortical surface visualizations of epileptiform activity. (A) Networks defined in the *DU15NET-Consensus Atlas*.^28^ The cortical surface overlap of voxels where nearby ictal (B), interictal (C), and ictal spread (D) activity were detected. DN-B = Default Network-B, FPN-B = Frontoparietal Network-B, SAL/PMN = Salience / Parietal Memory Network, CG-OP = Cingulo-Opercular, LANG = Language, DN-A = Default Network-A, FPN-A = Frontoparietal Network-A, AUD = Auditory, SMOT-B = Somatomotor-B, dATN-A = Dorsal Attention-A, dATN-B = Dorsal Attention-B, SMOT-A = Somatomotor-A, PM-PPr = Premotor-Posterior Parietal Rostral, VIS-P = Visual Peripheral, VIS-C = Visual Central.

The overall distributions of epileptiform activity type differed significantly across both regions and networks (all global Chi-squared tests *p* < 0.001). Follow-up testing revealed several specific anatomical regions and functional networks that exhibited significant deviations after correcting for multiple comparisons. For instance, all types of epileptiform activity were far more prevalent in temporal regions compared to the global baseline (Ictal: χ^2^ [1, n = 3683] = 216.61, Δ Observed-Expected [O-E] = +204.4%, *p* < 0.001; interictal χ^2^ [1, 3683] = 442.57, Δ O-E = +297.0%, *p* < 0.001; ictal spread χ^2^ [1, 3683] = 96.89, Δ O-E = +142.8%, *p* < 0.001). In contrast, frontal regions, which are densely sampled with iEEG, exhibited significantly less epileptiform activity than would be expected compared to the global baseline (Ictal: χ^2^ [1, 2047] = 109.86, Δ O-E = -65.4%, *p* < 0.001; interictal χ^2^ [1, 2047] = 232.43, Δ O-E = -75.8%, *p* < 0.001; ictal spread χ^2^ [1, 2047] = 89.17, Δ O-E = - 71.2%, *p* < 0.001) (**Figure 2B**).

Similarly, we observed multiple different networks that exhibited either significant increases or decreases in the proportion of epileptiform activity that would be expected. Notably, ictal and interictal activity was far more prevalent in DN-A (Default Network-A) (Ictal: χ^2^ [1, 646] = 22.32, Δ O-E = +84.6%, *p* < 0.001; interictal χ^2^ [1, 646] = 124.72, Δ O-E = +182.1%, *p* < 0.001), whereas interictal and ictal spread activity was more prevalent in VIS-P (Visual-Peripheral) network (Interictal: χ^2^ [1, 80] = 7.97, Δ O-E = +115.2%, *p* = 0.014; ictal spread χ^2^ [1, 80] = 7.06, Δ O-E = +141.0%, *p* = 0.019). All types of epileptiform activity were less prevalent in SAL/PMN (Salience / Parietal Memory Network) (Ictal: χ^2^ [1, 693] = 23.02, Δ O-E = -71.4%, *p* < 0.001; interictal χ^2^ [1, 693] = 33.37, Δ O-E = -72.4%, *p* < 0.001; ictal spread χ^2^ [1, 693] = 18.61, Δ O-E = -69.5%, *p* < 0.001) (**Figure 2C**). A summary of all Chi-squared tests is provided in **Table 2**.

**Table 2.** Summary of output from Chi-squared tests comparing the proportions of contacts exhibiting epileptiform activity across anatomical regions and functional networks. Rows are sorted by the total number of electrodes in each region/network. Δ O-E = Difference between the observed and expected proportion of electrodes with epileptiform activity, expressed as a percentage relative to the expected proportion. DN-B = Default Network-B, FPN-B = Frontoparietal Network-B, SAL/PMN = Salience / Parietal Memory Network, CG-OP = Cingulo-Opercular, LANG = Language, DN-A = Default Network-A, FPN-A = Frontoparietal Network-A, AUD = Auditory, SMOT-B = Somatomotor-B, dATN-A = Dorsal Attention-A, dATN-B = Dorsal Attention-B, SMOT-A = Somatomotor-A, PM-PPr = Premotor-Posterior Parietal Rostral, VIS-P = Visual Peripheral, VIS-C = Visual Central. Bolded *p\**-values indicate threshold for statistical significance (adjusted *p* < 0.05).

| Contrast | Ictal |  |  | Interictal |  |  | Ictal Spread |  |  |
| --- | --- | --- | --- | --- | --- | --- | --- | --- | --- |
| | $\chi^2$ | $\Delta$ O-E | $p^*$ | $\chi^2$ | $\Delta$ O-E | $p^*$ | $\chi^2$ | $\Delta$ O-E | $p^*$ |
| **Region** |  |  |  |  |  |  |  |  |  |
| Temporal | 216.61 | +204.4% | **< 0.001** | 442.57 | +297.0% | **< 0.001** | 96.89 | +142.8% | **< 0.001** |
| Frontal | 109.86 | -65.4% | **< 0.001** | 232.43 | -75.8% | **< 0.001** | 89.17 | -71.2% | **< 0.001** |
| Parietal | 0.19 | -6.6% | 0.784 | 20.25 | -46.8% | **< 0.001** | 7.09 | -42.1% | **< 0.001** |
| Subcortical | 34.51 | -81.4% | **< 0.001** | 51.87 | -82.2% | **< 0.001** | 7.18 | -47.7% | **0.019** |
| Insular | 13.93 | -62.2% | **0.001** | 7.72 | -38.9% | 0.015 | 0.58 | -17.8% | 0.579 |
| Occipital | 2.57 | -30.5% | 0.201 | 0.52 | -12.1% | 0.580 | 20.77 | +108.2% | **< 0.001** |
| **Network** |  |  |  |  |  |  |  |  |  |
| DN-B | 2.78 | -18.5% | 0.182 | 0.02 | -1.7% | 0.959 | 0.14 | +5.4% | 0.824 |
| FPN-B | 0.63 | +13.9% | 0.579 | 0.64 | -11.5% | 0.579 | 5.06 | -37.9% | 0.054 |
| SAL-PMN | 23.02 | -71.4% | **< 0.001** | 33.37 | -72.4% | **< 0.001** | 18.61 | -69.5% | **< 0.001** |
| CG-OP | 0.1 | +6.4% | 0.855 | 3.55 | -25.9% | 0.125 | 0.88 | +18.1% | 0.547 |
| LANG | 0.68 | +15.0% | 0.579 | 1.23 | -16.0% | 0.432 | 19.53 | +85.1% | **< 0.001** |
| DN-A | 22.32 | +84.6% | **< 0.001** | 124.72 | +182.1% | **< 0.001** | 0.67 | +16.2% | 0.579 |
| FPN-A | 0.00 | -2.8% | 0.967 | 14.09 | -55.7% | **0.001** | 8.27 | -55.0% | **0.012** |
| AUD | 0.71 | -24.1% | 0.579 | 2.79 | +38.3% | 0.182 | 5.02 | +65.7% | 0.054 |
| SMOT-B | 0.55 | -24.0% | 0.579 | 1.45 | +31.1% | 0.379 | 11.3 | -100.0% | **0.003** |
| dATN-A | 1.66 | +41.0% | 0.337 | 0.25 | +15.2% | 0.745 | 0.08 | +13.7% | 0.855 |
| dATN-B | 1.87 | -42.5% | 0.301 | 0.55 | -20.8% | 0.579 | 10.53 | +107.4% | **0.004** |
| SMOT-A | 0.58 | +32.1% | 0.579 | 3.38 | -56.6% | 0.134 | 0.01 | +10.1% | 0.963 |
| PM-PPr | 5.27 | +86.1% | 0.051 | 1.91 | +45.1% | 0.301 | 0.06 | -16.0% | 0.879 |
| VIS-P | 0.6 | +44.9% | 0.579 | 7.97 | +115.2% | **0.014** | 7.06 | +141.0% | **0.019** |
| VIS-C | 0.08 | -47.0% | 0.855 | 0.00 | -21.8% | 0.971 | 0.01 | -38.9% | 0.959 |

## Discussion

In this study, we systematically characterized the type and distribution of epileptiform activity across a large cohort of patients with drug-resistant epilepsy undergoing SEEG. Several key findings emerged. First, although all three forms of epileptiform activity—ictal, interictal, and ictal spread—were broadly observed across patients, their prevalence differed substantially, with interictal activity being the most common, followed by ictal activity and then ictal spread. Second, we found marked regional differences in the distribution of activity, with temporal regions frequently exhibiting epileptiform activity, whereas frontal regions showed significantly less activity than expected despite being densely sampled. Third, when considered within the framework of functional networks, we observed distinct deviations from baseline patterns, with some networks (e.g., DN-A, VIS-P) showing heightened epileptiform activity, and others (e.g., SAL/PMN) appearing less affected.

These results extend prior work by highlighting both the regional and network-level signatures of epileptiform activity in human epilepsy. The overrepresentation of activity within the temporal lobe is consistent with longstanding observations that mesial temporal lobe epilepsy represents the most common form of drug-resistant epilepsy.^17^ At the same time, the reduced prevalence of activity in frontal regions may reflect fundamental differences in the likelihood of observing pathological activity across brain regions. Importantly, our analyses at the network level reveal that epileptiform activity often localizes to networks involved in associative processing (e.g., DN-A), while other networks, such as salience and parietal memory systems, appear less susceptible.

The finding that interictal activity was the most common form of epileptiform activity, and that its distribution closely parallels that of ictal onsets, reinforces the clinical utility of interictal recordings as a surrogate biomarker for seizure localization. However, the presence of interictal activity in networks not traditionally associated with seizure onset, such as peripheral visual regions, raises important questions about whether these events represent truly epileptogenic signals, epiphenomena of broader network dysfunction, or secondary propagation. Further work integrating single-neuron recordings or scalp electroencephalography may help disentangle these possibilities.^32,33^

Finally, the network-level distribution of epileptiform activity observed here has potential translational implications. For surgical and neuromodulatory interventions, understanding the distributed architecture of epileptiform activity may help refine targeting strategies. For example, responsive neurostimulation may be particularly well suited for cases in which pathological activity is anchored in distributed networks, where focal resection is less feasible.^34^ Similarly, distinguishing between networks where epileptiform activity is widespread vs sparse could aid in predicting surgical outcomes and cognitive side effects, given the overlap between functional networks implicated in epilepsy and those supporting memory and higher cognition.^35^

### Limitations

Intracranial electrodes are generally placed in or near hypothesized seizure foci to aid in seizure localization. This presents two limitations for the present study: First, this approach results in a tendency to oversample areas frequently implicated in epileptogenesis (e.g., mesial temporal lobe). Subsequent analyses may, therefore, be biased by both the paucity of recordings in other regions and the dense spatial proximity of electrodes within areas that are oversampled—neighboring leads are likely to detect the same current source, which may artificially inflate the proportion of electrodes that exhibited epileptiform activity. Second, sampling of a less common area (e.g., occipital lobe) is likely motivated by a strong hypothesis that the region is pathological, whether based on seizure semiology or prior investigations. Thus, estimates of the prevalence of epileptiform activity in infrequently sampled regions are likely to be more variable than those with more recordings. To minimize bias in our methodological approach, we used analyses that controlled for differences in the number of electrodes across recorded regions (i.e., Chi-squared tests).

Nonetheless, given these limitations, the results of the study should be considered as descriptive, rather than as providing probabilistic claims regarding the potential yield of sampling different anatomical regions or functional networks with intracranial recordings. Moreover, it would be difficult, in practice, to target specific functional networks with existing linear SEEG leads since they involve distributed cortical regions rather than a spatially contiguous region.

### Conclusions

In summary, this study provides a comprehensive, large-scale characterization of epileptiform activity across both anatomical regions and functional networks in patients with drug-resistant epilepsy. Our findings support the view that epilepsy reflects both focal pathology and network-level dysfunction, which has important implications for surgical planning, biomarker development, and the design of future neuromodulatory therapies. By mapping epileptiform activity within a network framework, our work contributes to a growing understanding of epilepsy as a disorder of distributed brain systems and lays the foundation for more precise, circuit-based interventions.

## Supporting information

Supplemental Authorship and Acknowledgments

## Data Availability

University of Utah Hospital

