## Supplemental Authorship and Acknowledgments for "Intracranial-EEG Based Classification and Localization of Epileptiform Activity in a Large Cohort of Adults with Drug-Resistant Epilepsy"

### Supplemental Information

*Keywords*

- Epilepsy
- Ictal
- Interictal
- SEEG
- Network

*Key Points*

- Interictal activity was common, detected in ~10% of contacts per patient
- Epileptiform activity was prevalent in temporal regions and reduced in frontal regions
- Associative functional networks exhibited increased epileptiform activity

*Acknowledgments*

We thank Kristin Krauss for her editorial assistance.

*Funding*

None.

*Conflict of Interest Statement*

The authors have no conflicts of interest to declare.

*Ethics Approval Statement*

This retrospective review was approved by the University of Utah Institutional Review Board (IRB # ***) with a waiver of patient consent.

*Data Availability Statement*

Data will not be made publicly available to protect patient privacy. Further inquiries can be directed to the corresponding author.

*Author Contributions (CRediT Taxonomy)*

- SY: Conceptualization, Data Curation, Writing – Review & Editing
- JC: Conceptualization, Formal Analysis, Visualization, Software, Data Curation, Writing – Original Draft
- MF: Conceptualization, Data Curation, Writing – Review & Editing
- WS: Formal Analysis, Visualization, Software, Data Curation, Writing – Review & Editing
- KM: Data Curation, Writing – Review & Editing
- SS: Data Curation, Writing – Review & Editing
- CS: Data Curation, Writing – Review & Editing
- AA: Investigation, Resources, Writing – Review & Editing
- AP: Investigation, Resources, Writing – Review & Editing
- SR: Investigation, Resources, Writing – Review & Editing
- BN: Investigation, Resources, Writing – Review & Editing
- BJ: Investigation, Resources, Writing – Review & Editing
- MJ: Investigation, Resources, Writing – Review & Editing
- KK: Investigation, Resources, Writing – Review & Editing
- SR: Resources, Writing – Review & Editing

BS: Conceptualization, Project Administration, Supervision, Resources, Writing – Review & Editing
